# A novel parameter for predicting immediate postoperative shoulder balance in Lenke Type 1 and 2 adolescent idiopathic scoliosis patients

**DOI:** 10.64898/2026.01.26.26344281

**Authors:** Pengfei Chi, Zheng Tian, Bin Zhang, Wen Yu, Junyao Cheng, Kaige Mao, Zheng Wang, Kai Song

## Abstract

**BACKGROUND CONTEXT:** Postoperative shoulder imbalance (PSI) is common following posterior spinal fusion (PSF) surgery in Lenke Type 1 and 2 adolescent idiopathic scoliosis (AIS) patients. However, it remains unclear that why preoperative shoulder balance cannot predict postoperative shoulder balance (PSB).

**PURPOSE:** To evaluate the predictive value of the thoracic cage-clavicle angle (TCCA) for immediate PSB.

**STUDY DESIGN/SETTING:** Retrospective cross-sectional study

**METHODS:** The degree of clavicle angle (CA), thoracic cage tilt angle (TCTA), thoracic spine tilt angle (TSTA), proximal thoracic curve (PTC) Cobb angle, and main thoracic curve (MTC) Cobb angle were measured before and after surgery. Six parameters, including TCCA, thoracic spine-clavicle angle (TSCA), CCAD-based CA (CCAD-CA), correction rate of PTC, correction rate of MTC, relative PT/MT residual Cobb angle (RRCA), were calculated. Intra-class correlation coefficient (ICC) analysis was performed for TCTA. Multinomial logistic regression was used to determine the risk/protective factors of PSB. A *p*-value of less than 0.05 was considered statistically significant.

**RESULTS:** The intra-rater ICCs for TCTA were 0.974, 0.946, and the inter-rater ICC was 0.887 before surgery. The intra-rater ICCs for TCTA were 0.975, 0.938, and the inter-rater ICC was 0.826 after surgery. For TCCA, in R group (*vs*. B group), pre-op right shoulder high (TCCA>0) and RRCA were risk factors. Pre-op left shoulder high (TCCA<0) and correction rate of MTC were protective factors. In L group (*vs*. B group), pre-op left shoulder high (TCCA<0) was a risk factor. Pre-op right shoulder high (TCCA>0) was a protective factor.

**CONCLUSIONS:** In Lenke Type 1 and 2 AIS patients, TCCA-based preoperative shoulder balance serves as a direction-specific predictor for immediate PSB. It acts as a significant risk factor for the postoperative ipsilateral shoulder high, while conversely functioning as a significant protective factor against the postoperative contralateral shoulder high.

## Introduction

Shoulder balance plays a significant role in the overall cosmetic appearance among patients with adolescent idiopathic scoliosis (AIS)[1]. However, postoperative shoulder imbalance (PSI) is common following posterior spinal fusion (PSF) surgery[2]. PSI could indicate surgical decompensation, the need for possible reoperation, and other complications, such as distal adding-on phenomenon, new lumbar curve (LC) progression, and trunk shift[3].

Many previous studies have indicated that preoperative shoulder status cannot predict postoperative shoulder balance (PSB)[2, 4-6]. This is a puzzling phenomenon, and we believe that it likely indicates some overlooked factors. Prior studies of preoperative shoulder balance have predominantly employed the horizontal plane as the reference frame. While in clinical practice, we have observed that Lenke Type 1 and 2 AIS patients are frequently accompanied by thoracic cage tilt in coronal plane, which appears to improve postoperatively.

Given the anatomical relationship between the thoracic cage and the shoulder, we speculate that assessment of preoperative shoulder balance using the tilted thoracic cage as a reference frame, rather than the horizontal plane, may improve the prediction of PSB. Therefore, we propose the concept of the thoracic cage-clavicle angle (TCCA). The aim of this study is to evaluate its predictive value for immediate PSB.

## Materials and Methods

### Patient recruitment

This was a retrospective study. Lenke Type 1 and 2 AIS patients who received posterior spinal fusion surgery in our institution from 2013 to 2024 were recruited in the study. The inclusion criteria were as follows: a diagnosis of adolescent idiopathic scoliosis, posterior spinal fusion surgery, complete standing full-length posteroanterior spine radiographs taken before surgery (pre-op), and immediately after surgery (post-op). All radiographs were acquired with the patients in a natural standing position, with their knees fully extended and arms naturally positioned at the sides. Immediate postoperative radiographs were taken when the patient was able to stand naturally after surgery. The exclusion criteria were as follows: previous surgery to the spine, pelvis, or lower limbs, other developmental abnormalities of the pelvis and lower limbs, scoliosis due to congenital, neuromuscular, syndromic, or etiologies other than AIS, radiographs without clavicle and humeral head. The demographic data were recorded including age at surgery, gender, and curve type by Lenke classification.

### Radiographic Measurements

The coronal parameters were measured on standing full-length posteroanterior spine radiographs that were taken before and after surgery, including (1) Clavicle angle (CA), the angle between the horizontal line and the line connecting the highest points of each clavicle, with the right side higher defined as positive, (2) Thoracic cage tilt angle (TCTA), the average value of the two angles between the vertical line and the line connecting the most lateral points of the 4th and 8th ribs, with a leftward inclination of the upper thoracic cage defined as positive (Fig 1a), (3) Thoracic spine tilt angle (TSTA), the angle between the vertical line and the line connecting the centroid of T1 and the centroid of T12, with a leftward deviation of the centroid of T1 defined as positive (Fig 1b), (4) Proximal thoracic curve (PTC) Cobb angle, (5) Main thoracic curve (MTC) Cobb angle. All parameters were measured independently by two senior spine surgeons, and the mean values were calculated for further analysis. Six parameters were calculated as follows: (1) TCCA (°) = CA-TCTA (TCCA > 0, right shoulder high; TCCA < 0, left shoulder high) (Fig 1a), (2) thoracic spine-clavicle angle(TSCA, °) = CA-TSTA (TSCA > 0, right shoulder high; TSCA < 0, left shoulder high) (Fig 1b), (3) CCAD-CA (°) = right clavicle chest cage angle (RCCA) - left clavicle chest cage angle (LCCA)[7] (CCAD-CA > 0, right shoulder high; CCAD-CA < 0, left shoulder high) (Fig 1c), (4) Correction rate of PTC= (pre-op PTC Cobb - post-op PTC Cobb)/pre-op PTC Cobb×100%, (5) Correction rate of MTC= (pre-op MTC Cobb - post-op MTC Cobb)/pre-op MTC Cobb×100%, (6) Relative PT/MT Residual Cobb angle (RRCA) = post-op PTC Cobb/ post-op MTC Cobb×100%.

**Fig. 1.**
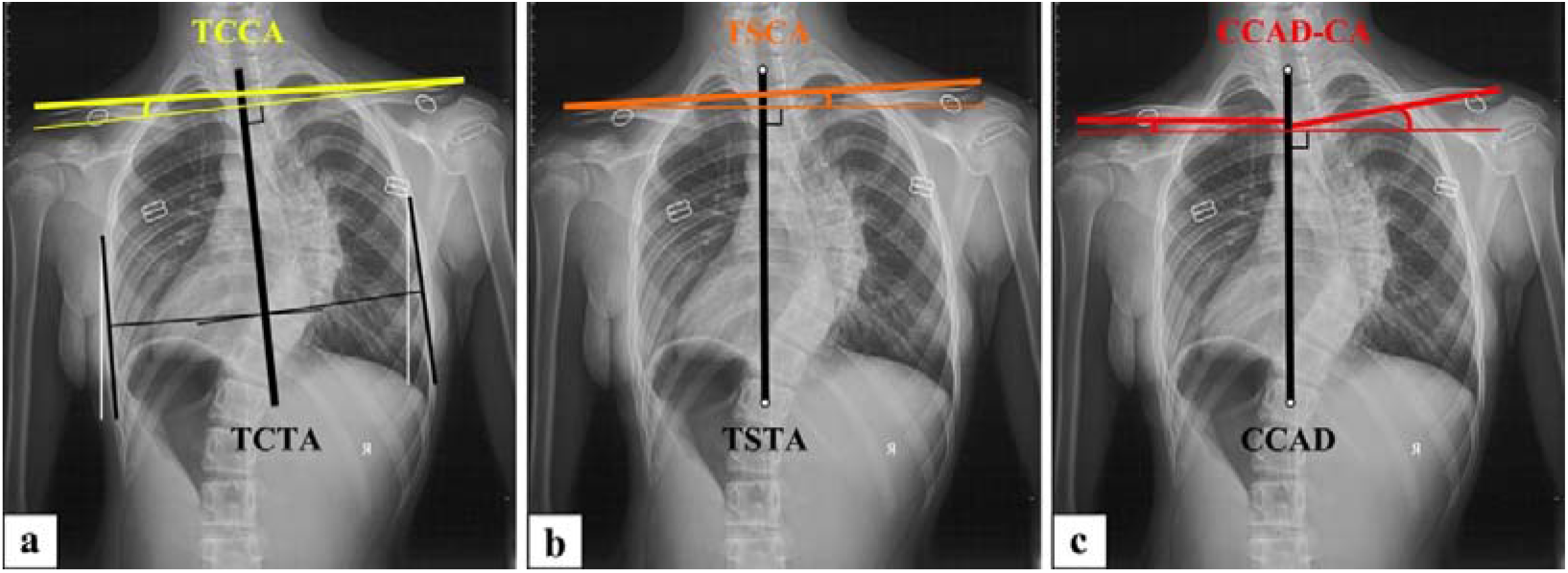
Radiographs of a girl in her 10’s with AIS. **a** Preoperative TCTA=6.20°, TCCA=-2.84°. **b** Preoperative TSTA=0.39°, TSCA=2.97°. **c** Preoperative CCAD-CA=9.70°.

PSI was defined as a postoperative absolute clavicle angle (CA) value of > 2°[1]. The patients were divided into three groups on the basis of postoperative CA: (1) postoperative right shoulder high group (R group): postoperative CA > 2°, (2) postoperative left shoulder high group (L group): postoperative CA < -2°, and (3) postoperative balanced group (B group): -2° ≤ postoperative CA ≤ 2°.

### Reliability Evaluation

Intraclass correlation coefficient (ICC) analysis with the corresponding 95% confidence interval (CI) was performed for TCTA to determine its intra- and inter-rater reliability. Two senior spine surgeons independently measured TCTA twice at an interval of 2 weeks. ICCs were based on a mean-rating (*k*=2), absolute-agreement, two-way random-effects model. ICC was considered to be excellent (ICC >0.90), good (ICC 0.75-0.90), moderate (ICC 0.50-0.75) or poor (ICC <0.50)[8].

### Statistical analysis

The coronal parameters were statistically analyzed. A paired *t*-test was used to compare the differences in the parameters before and after surgery. Multinomial logistic regression was used to determine the risk factors of PSI. B group was categorized as reference category. The model was adjusted for potential confounders including CA-based preoperative shoulder balance, correction rate of PTC, correction rate of MTC, RRCA, and upper instrumented vertebra (UIV). The likelihood ratio test was used to assess overall model fit and variable significance. Results were presented as odds ratios (ORs) with 95% CIs. Multiple linear regression analysis was performed to predict post-op CA based on the correction degree of PTC and MTC. *p* < 0.05 indicated a statistically significant difference. Statistical software SPSS 22.0 was used for data analysis.

## Results

There were 154 patients (aged from 11 to 19 years) in the study population. The demographics and radiographic parameters are listed in Table 1.

**Table 1.**
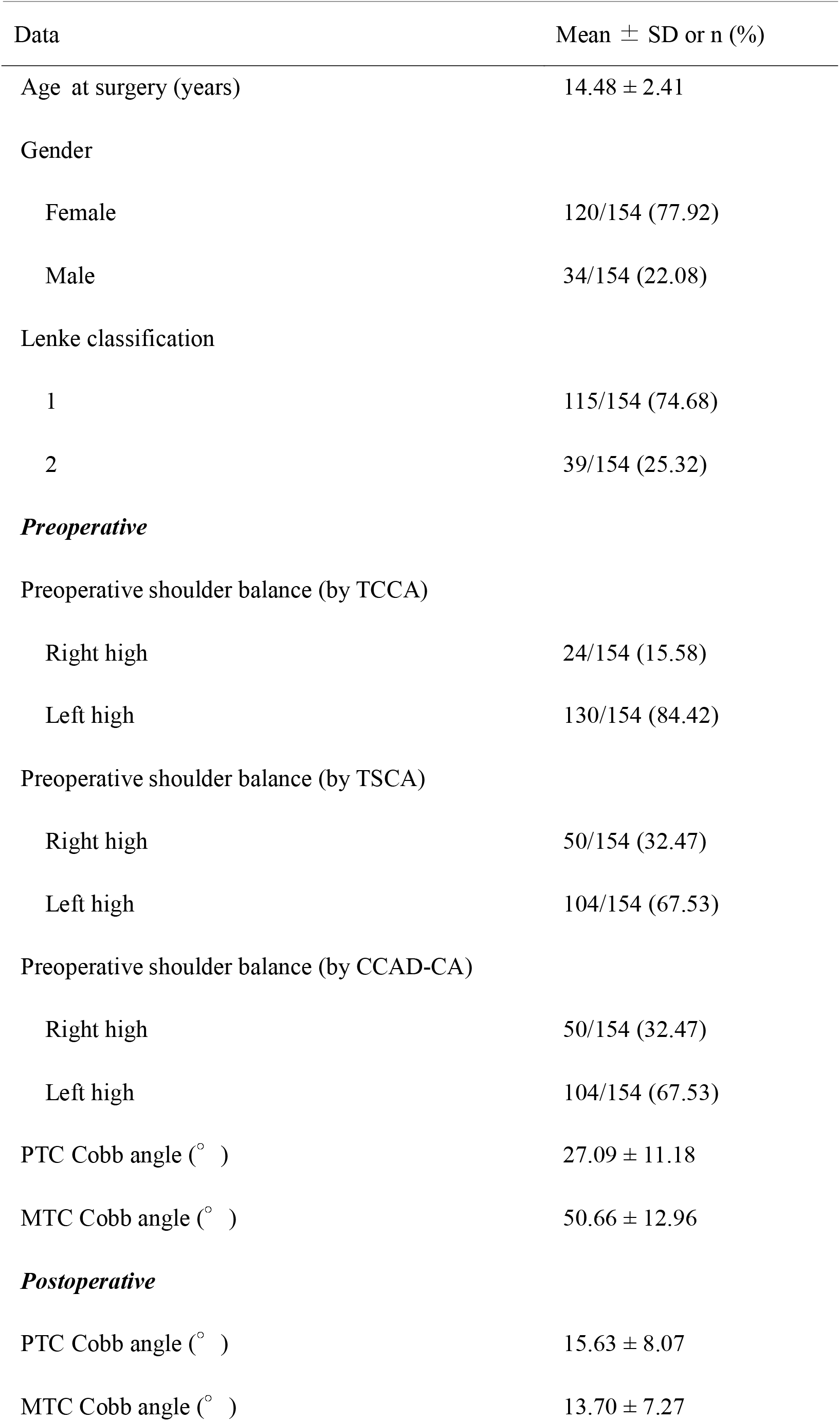
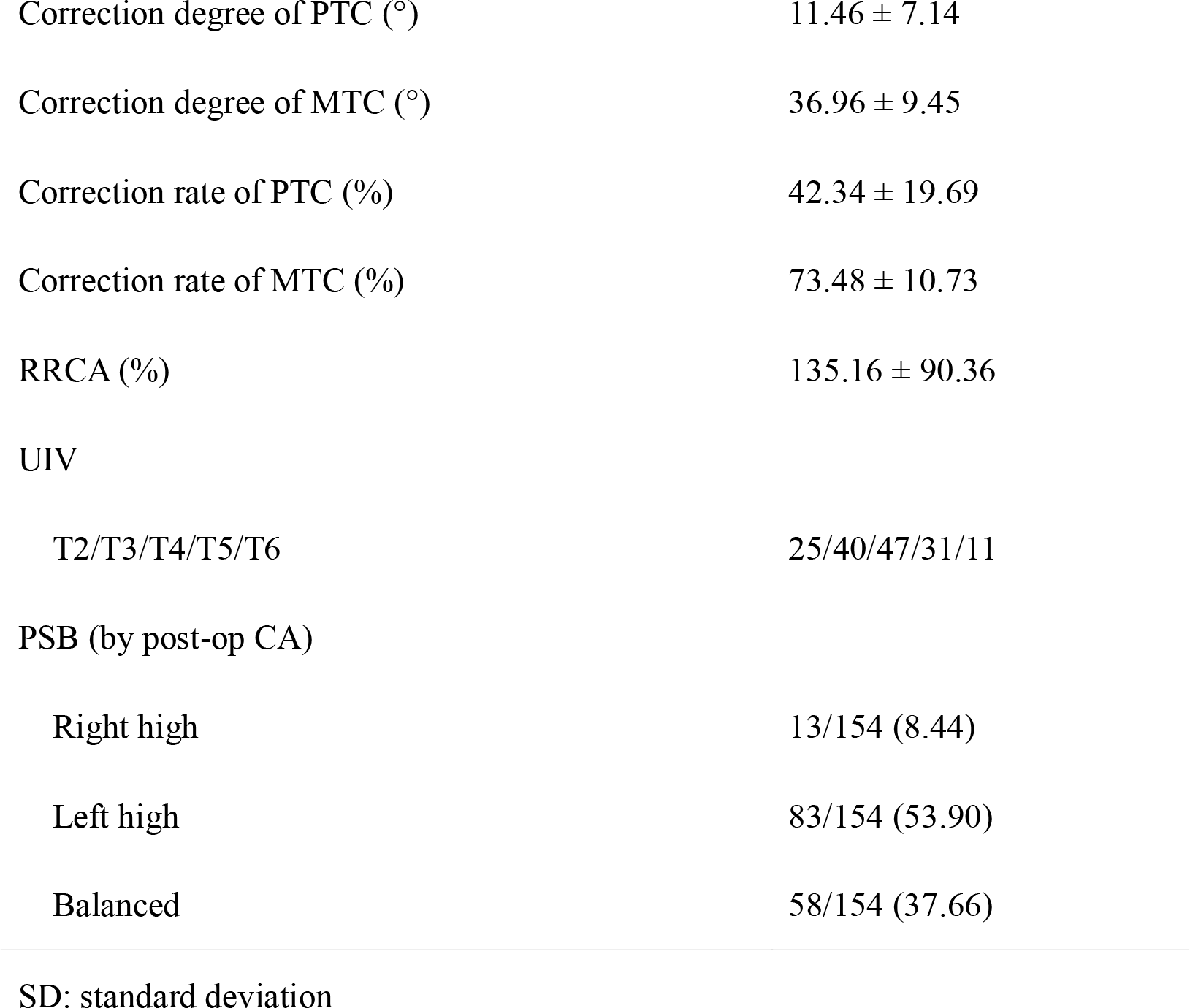
Demographic data and radiographic parameters of Lenke Type 1 and 2 AIS.

| Data | Mean ± SD or n (%) |
| --- | --- |
| Age at surgery (years) | 14.48 ± 2.41 |
| Gender |  |
| Female | 120/154 (77.92) |
| Male | 34/154 (22.08) |
| Lenke classification |  |
| 1 | 115/154 (74.68) |
| 2 | 39/154 (25.32) |
| ***Preoperative*** |  |
| Preoperative shoulder balance (by TCCA) |  |
| Right high | 24/154 (15.58) |
| Left high | 130/154 (84.42) |
| Preoperative shoulder balance (by TSCA) |  |
| Right high | 50/154 (32.47) |
| Left high | 104/154 (67.53) |
| Preoperative shoulder balance (by CCAD-CA) |  |
| Right high | 50/154 (32.47) |
| Left high | 104/154 (67.53) |
| PTC Cobb angle (° ) | 27.09 ± 11.18 |
| MTC Cobb angle (° ) | 50.66 ± 12.96 |
| ***Postoperative*** |  |
| PTC Cobb angle (° ) | 15.63 ± 8.07 |
| MTC Cobb angle (° ) | 13.70 ± 7.27 |
| Correction degree of PTC (°) | 11.46 ± 7.14 |
| Correction degree of MTC (°) | 36.96 ± 9.45 |
| Correction rate of PTC (%) | 42.34 ± 19.69 |
| Correction rate of MTC (%) | 73.48 ± 10.73 |
| RRCA (%) | 135.16 ± 90.36 |
| UIV |  |
| T2/T3/T4/T5/T6 | 25/40/47/31/11 |
| PSB (by post-op CA) |  |
| Right high | 13/154 (8.44) |
| Left high | 83/154 (53.90) |
| Balanced | 58/154 (37.66) |

SD: standard deviation

### Reliability of TCTA

Before surgery, the intra-rater ICC for TCTA can be regarded as “excellent” (0.974 and 0.946). The inter-rater ICC was “good” (0.887). After surgery, the intra-rater ICC for TCTA was “excellent” (0.975 and 0.938). The inter-rater ICC was “good” (0.826). Theoretically, when measuring TCTA, symmetry in the starting and ending points of the rib tangents on both sides is sufficient. However, considering that the mid-thoracic region is relatively flat and easier to measure, it is recommended that the region of the ribs range from the 4th to the 8th ribs on both sides.

The comparisons of coronal parameters associated with CA in Lenke Type 1 and 2 AIS patients are presented in Table 2. Significant differences were observed in CA, TCTA, TSTA, TCCA, TSCA, and CCAD-CA before and after surgery.

**Table 2.**
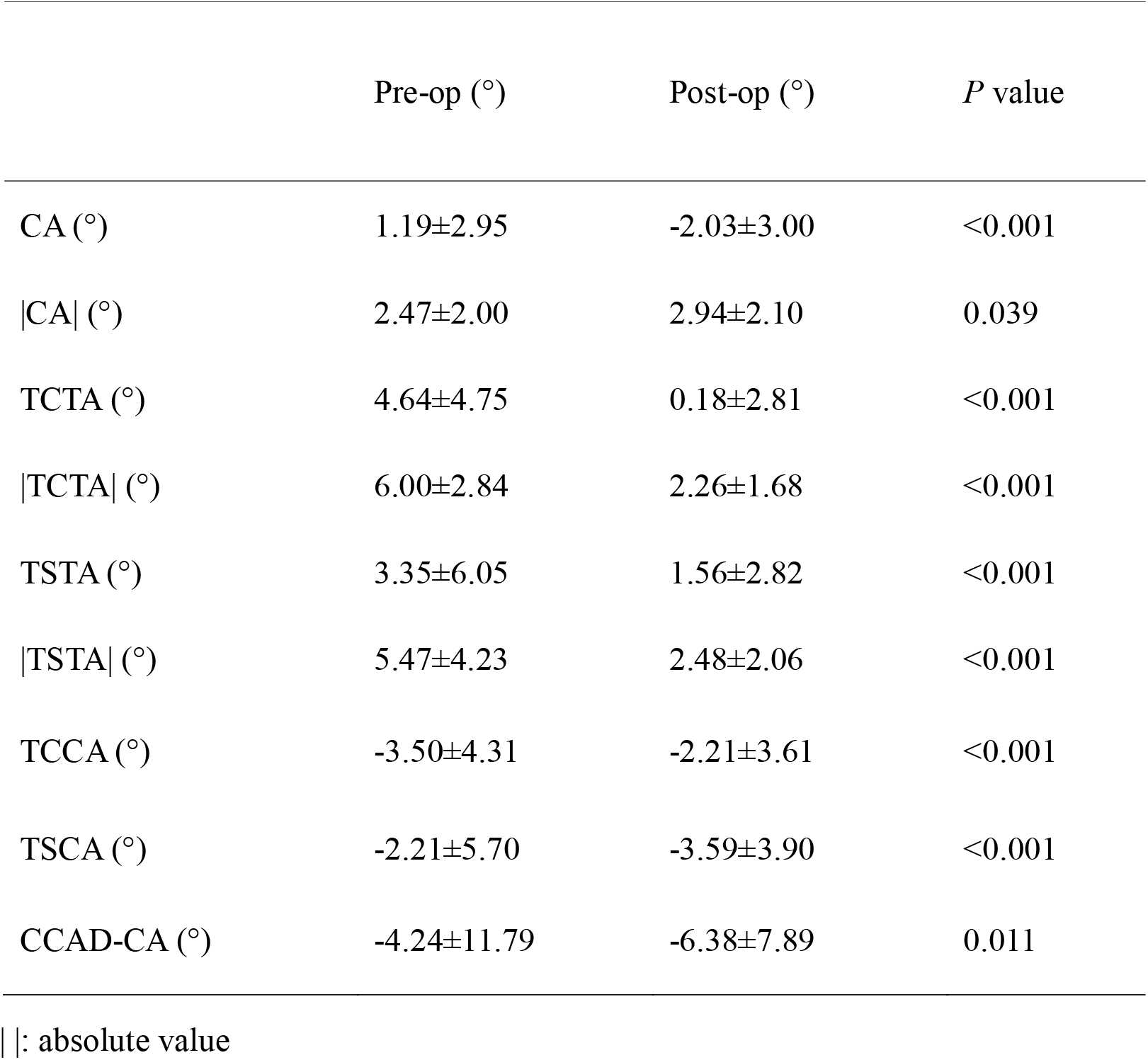
Comparisons of coronal parameters associated with CA before and after surgery.

### Likelihood ratio tests

Likelihood ratio tests for the Multinomial logistic regression model are presented in Table 3. In terms of TCCA, TCCA-based preoperative shoulder balance (left/right high), correction rate of MTC, RRCA were statistically significantly associated with PSB, while CA-based preoperative shoulder balance (left/right high), correction rate of PTC and UIV did not show the significant association.

**Table 3.** Likelihood ratio tests for the multinomial logistic regression model.

| Effect | $\chi^2$ | df | *P* value |
| --- | --- | --- | --- |
| ***TCCA*** |  |  |  |
| Overall model | 53.614 | 18 | <0.001 |
| Preoperative shoulder balance (by TCCA) | 27.342 | 2 | <0.001 |
| Preoperative shoulder balance (by CA) | 1.500 | 2 | 0.472 |
| Correction rate of PTC | 2.109 | 2 | 0.348 |
| Correction rate of MTC | 9.202 | 2 | 0.010 |
| RRCA | 8.249 | 2 | 0.016 |
| UIV | 9.358 | 8 | 0.313 |
| ***TSCA*** |  |  |  |
| Overall model | 36.099 | 18 | 0.007 |
| Preoperative shoulder balance (by TSCA) | 9.826 | 2 | 0.007 |
| Preoperative shoulder balance (by CA) | 0.743 | 2 | 0.690 |
| Correction rate of PTC | 2.733 | 2 | 0.255 |
| Correction rate of MTC | 7.567 | 2 | 0.023 |
| RRCA | 6.709 | 2 | 0.035 |
| UIV | 9.923 | 8 | 0.270 |
| ***CCAD-CA*** |  |  |  |
| Overall model | 37.044 | 18 | 0.005 |
| Preoperative shoulder balance (by CCAD-CA) | 10.771 | 2 | 0.005 |
| Preoperative shoulder balance (by CA) | 0.650 | 2 | 0.722 |
| Correction rate of PTC | 2.294 | 2 | 0.318 |
| Correction rate of MTC | 6.184 | 2 | 0.045 |
| RRCA | 6.570 | 2 | 0.037 |
| UIV | 9.833 | 8 | 0.277 |

In terms of TSCA, TSCA-based preoperative shoulder balance (left/right high), correction rate of MTC, and RRCA were statistically significantly associated with PSB, while CA-based preoperative shoulder balance (left/right high), correction rate of PTC, and UIV did not show the significant association.

In terms of CCAD-CA, CCAD-CA-based preoperative shoulder balance (left/right high), correction rate of MTC, and RRCA were statistically significantly associated with PSB, while CCAD-CA-based preoperative shoulder balance (left/right high), correction rate of PTC, and UIV did not show the significant association.

### Multinomial logistic regression analysis

Multinomial logistic regression analysis of factors associated with PSB are showed in Table 4. For TCCA, in R group (*vs*. B group), pre-op right shoulder high (TCCA>0) and RRCA were risk factors. Pre-op left shoulder high (TCCA<0) and correction rate of MTC were protective factors. CA-based preoperative shoulder balance, correction rate of PTC showed no significant effect.

**Table 4.**
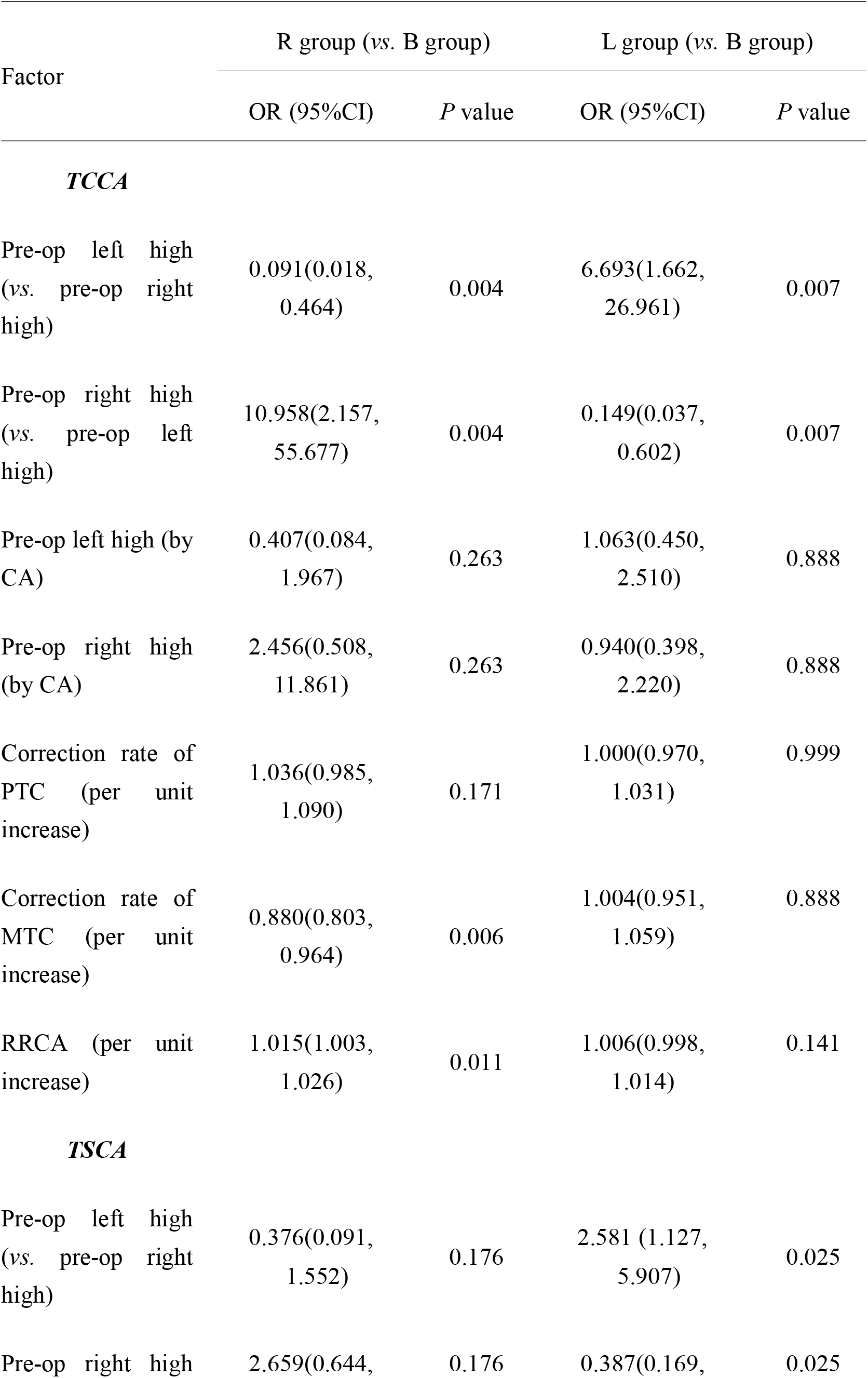
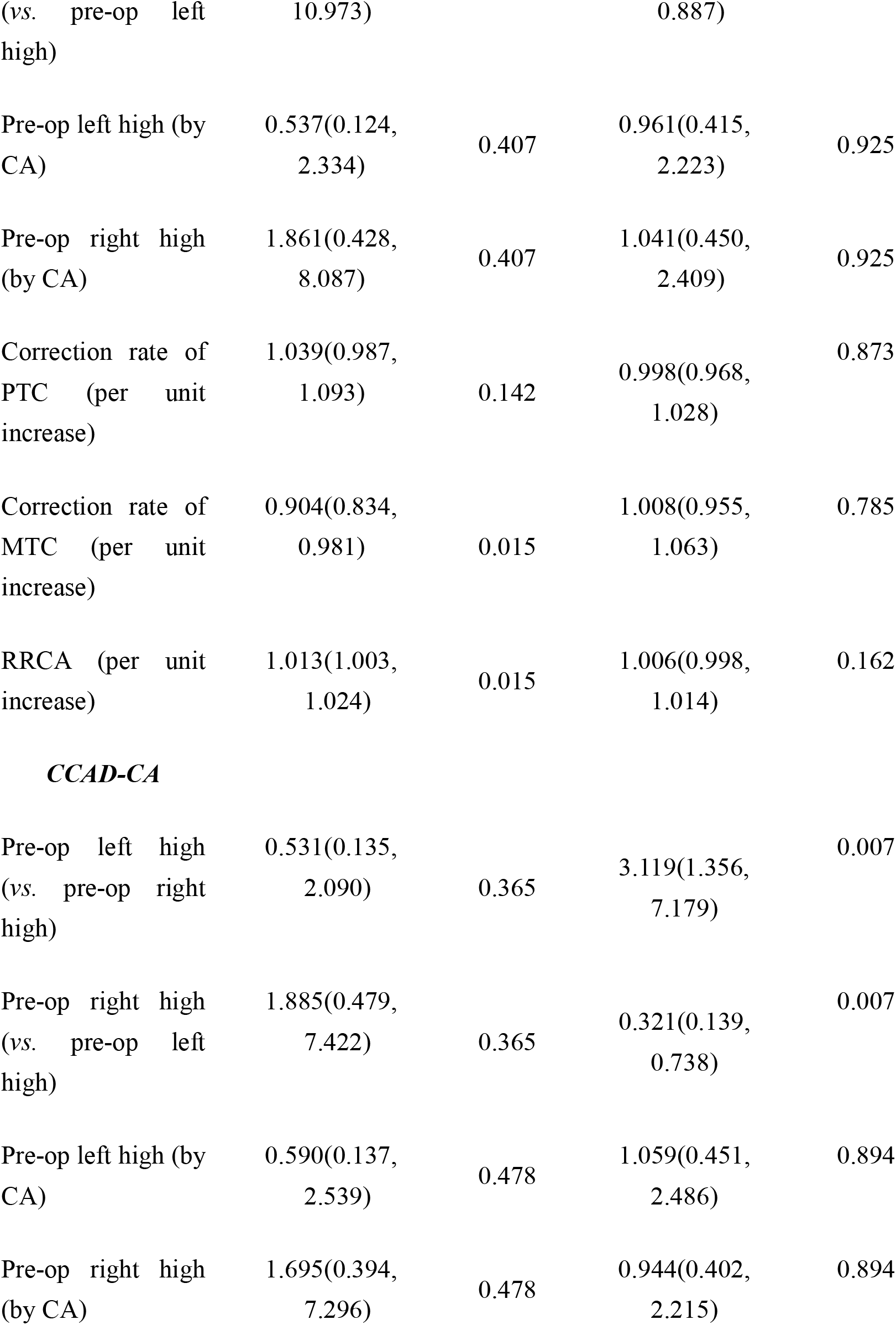
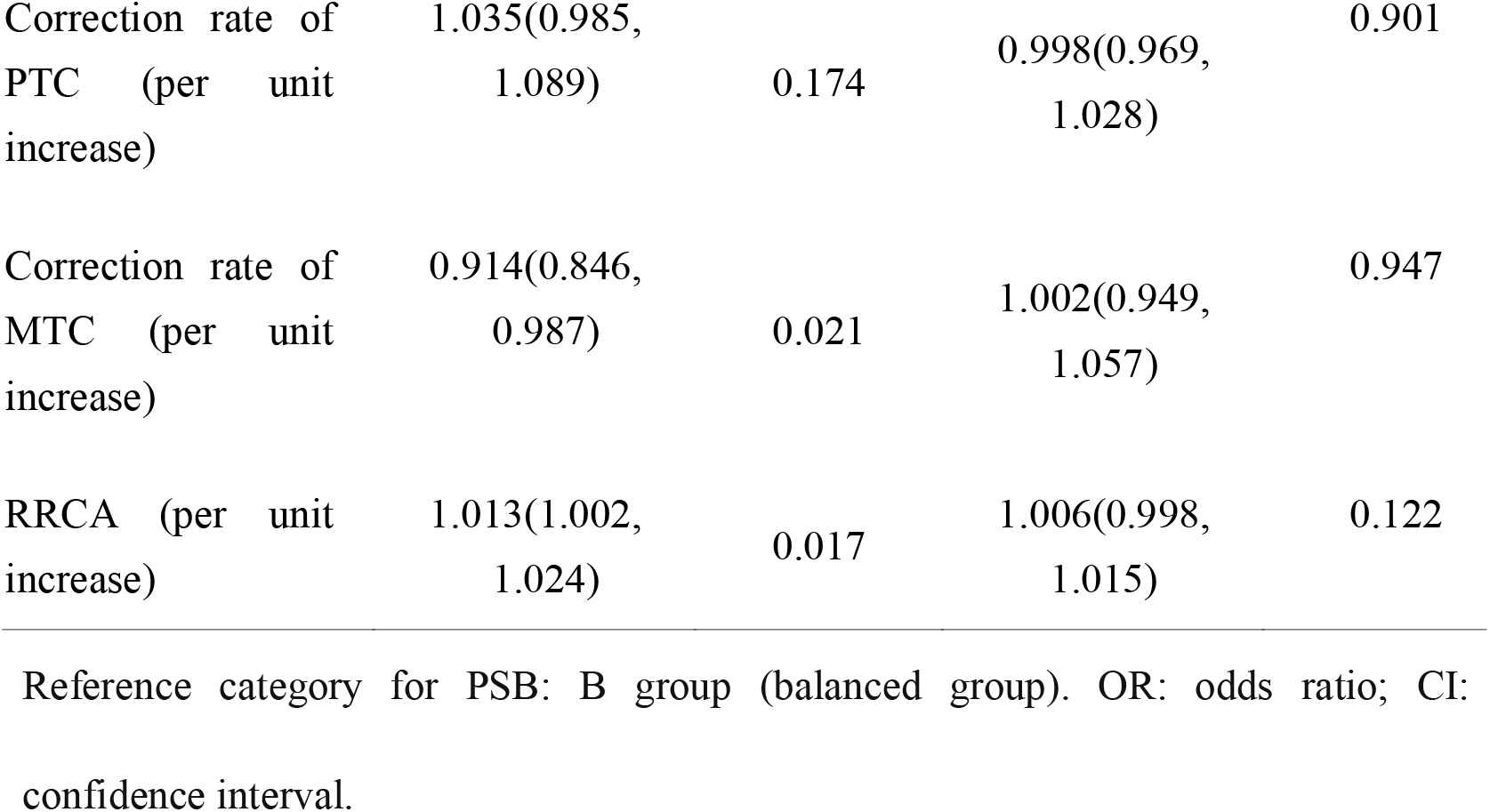
Multinomial logistic regression analysis of factors associated with PSB.

In L group (*vs*. B group), pre-op left shoulder high (TCCA<0) was a risk factor. Pre-op right shoulder high (TCCA>0) was a protective factor. CA-based preoperative shoulder balance, correction rate of PTC, correction rate of MTC, and RRCA showed no significant effect.

For TSCA, in R group (*vs*. B group), RRCA was a risk factor. Correction rate of MTC was a protective factor. TSCA-based and CA-based pre-op shoulder balance, and correction rate of PTC showed no significant effect.

In L group (*vs*. B group),pre-op left shoulder high (TSCA<0) was a risk factor. Pre-op right shoulder high (TSCA>0) was a protective factor. CA-based preoperative shoulder balance, correction rate of PTC, correction rate of MTC, and RRCA showed no significant effect.

For CCAD-CA, in R group (*vs*. B group), RRCA was a risk factor. Correction rate of MTC was a protective factor. CCAD-CA-based and CA-based pre-op shoulder balance, and correction rate of PTC showed no significant effect.

In L group (*vs*. B group), pre-op left shoulder high (CCAD-CA<0) was a risk factor. Pre-op right shoulder high (CCAD-CA>0) was a protective factor. CA-based pre-op shoulder balance, correction rate of PTC, correction rate of MTC, and RRCA showed no significant effect.

Based on the comparison of the ORs with 95% CIs of TCCA/TSCA/CCAD-CA related parameters, we recommend using TCCA to predict immediate PSB.

### Multiple linear regression analysis

#### The predictive equation was as follows

Post-op CA = pre-op TCCA + (0.22 × correction degree of PTC) - (0.10 × correction degree of MTC) + 2.86 (R^2^=0.127; correction degree of PTC: B=0.22, *p*<0.001; correction degree of MTC: B=-0.10, *p*=0.009) ⍰

## Discussion

### Predictive value of TCCA

In clinical practice, we observed that Lenke type 1 and 2 AIS patients frequently presented with thoracic cage tilt in coronal plane, and such tilt tended to improve after surgery. Considering the anatomical relationship between the thoracic cage and the clavicle, TCCA was proposed to predict immediate PSB.

We believe that the inability of the conventional preoperative CA to predict PSB may stem from the fact that the conventional CA incorporates the influence of thoracic cage tilt, and it is not the “true CA”. In contrast, TCCA eliminates the effect of thoracic cage tilt, thereby representing the “true CA”. This may explain the predictive value of TCCA for PSB.

How to understand the role of TCCA? In practical application, the role of TCCA is equivalent to rotating the preoperative full-length spinal radiograph in coronal plane to align the thoracic cage perpendicular to the horizontal line. This adjustment eliminates the influence of preoperative thoracic cage tilt on shoulder balance, thereby revealing the “true CA”. This allows for a preliminary assessment of preoperative shoulder balance and PSB (Fig. 2). Ultimately, however, the calculation of TCCA is necessary to formulate the surgical strategy before surgery.

**Fig. 2.**
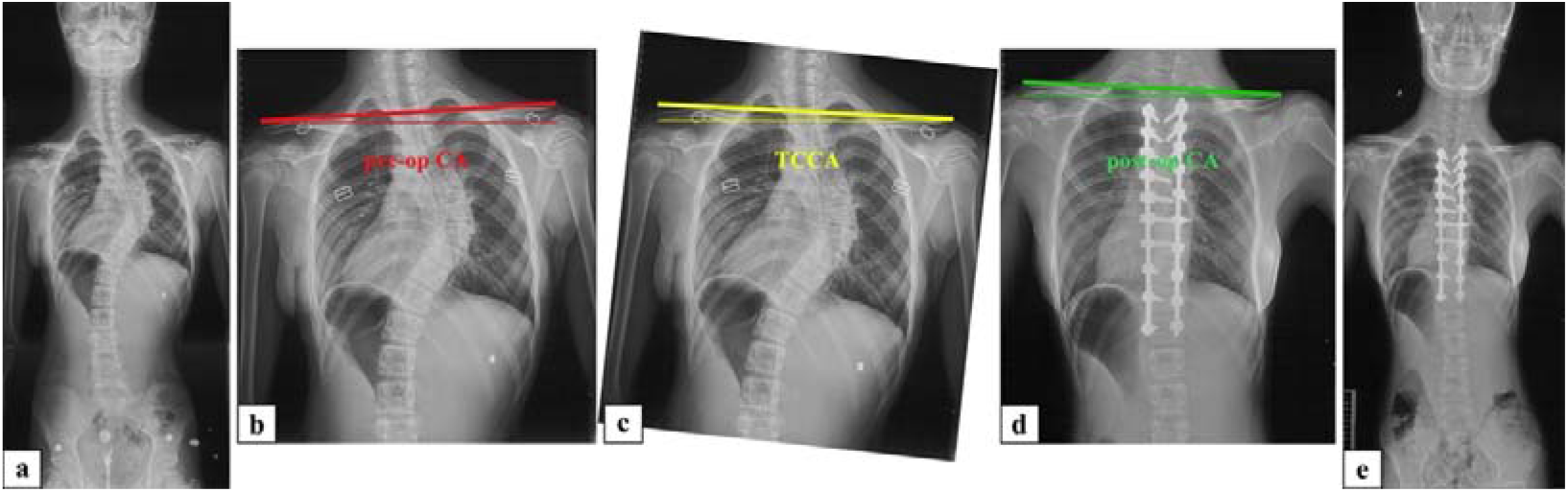
Radiographs of a girl in her 10’s with AIS. **a** AIS with thoracic cage tilt in coronal plane before surgery. **b** Preoperative CA=3.36° (right shoulder high). **c** Rotate the radiograph to orient the thoracic cage perpendicular to the horizontal line, TCCA=-2.84° (left shoulder high). **d** Postoperative CA=-2.76 ° (left shoulder high). **e** Thoracic cage tilt improved after surgery.

The superior predictive ability of TCCA over TSCA and CCAD-CA for PSB may be attributed to the better improvement in thoracic cage tilt. Although TCCA, TSCA, and CCAD-CA showed significant changes after surgery, their mean values remained negative, which had little impact on their assessments of shoulder balance. Furthermore, given the correction of TSTA was not as substantial as that of TCTA (1.56° *vs*. 0.18°), the predictive reliability of TSCA and CCAD-CA was limited.

Sielatycki et al.[9] recommend instrumenting to T2 if the left shoulder is high preoperatively, T3 if the shoulders are balance, and T4 if the left shoulder is down in a right-thoracic Lenke Type 1 or 2 AIS patient for ensuring PSB. However, numerous previous studies have consistently shown no significant association between preoperative and postoperative shoulder balance. We speculate that this may be because previous studies employed the horizontal plane as the reference frame, thereby overlooking the confounding effect of the thoracic cage. As a result, we propose modifying Sielatycki’s surgical strategies by incorporating TCCA-based preoperative shoulder balance, which may offer greater clinical relevance.

Regarding the CCAD, it is important to note that Yagi’s study[7] used the magnitude of the CCAD to predict PSI and our approach modified this concept. We utilized the CCAD to estimate the theoretical preoperative left-right shoulder height (CCAD-CA > 0, right shoulder high; CCAD-CA < 0, left shoulder high), which differed from the original methodology. This adaptation was made to facilitate comparison with the other two parameters. Our findings indicated that the theoretical preoperative shoulder balance based on CCAD-CA had a lower predictive value for PSB compared to TCCA and TSCA. Although, under ideal conditions, the CCAD-CA-based preoperative shoulder balance should align with that measured by TSCA, factors such as rotation and differences in the relative vertical positions of the bilateral clavicles resulted in discrepancies between the theoretical CCAD-CA-based shoulder balance and the actual anatomical shoulder balance. Consequently, the CCAD-CA demonstrated limited efficacy in predicting PSB.

### Other confounding factors

As this study focused on immediate PSB rather than long-term follow-up data, parameters directly related to the surgical procedure were selected for analysis. Previous studies have shown that the correction rate of PTC, correction rate of MTC, and RRCA were risk factors for PSI [1, 2, 6, 10, 11].

Farshad et al.[3] found that preoperative level shoulder was a risk factor for PSI and Tomohiro et al.[11] reported that preoperative lower right shoulder elevation had a higher risk of PSI when the MT curve was well corrected. These may be attributed to the fact that since right-sided thoracic curves predominate in AIS patients, the proximal thoracic cage tends to tilt to the left. Preoperative level shoulder or even slightly elevated right shoulder may actually demonstrate left shoulder high as measured by TCCA. If these patients are erroneously treated based on the appearance of right shoulder high, intentionally increasing the correction rate of MTC intraoperatively may result in overcorrection and ultimately exacerbate left shoulder elevation postoperatively.

Many previous studies reported that insufficient correction of PTC and overcorrection of MTC showed high risk of developing PSI in AIS patients[2, 4, 5, 9, 11, 12]. However, Gotfryd et al.[13] reported no correlation between PSB and the amount of PT and MT curve correction for Lenke Type 1. Chan et al.[1] found that in Lenke Type 1 and 2 patients, RRCA had shown a significant correlation with medial shoulder balance, and Lee et al.[6] reported that in Lenke Type 2 patients, a higher postoperative PTC/MTC ratio was a risk factor for PSI.

Although, in our study, the correction rate of PTC showed no significant effect on PSB, and RRCA was an independent risk factor for postoperative right shoulder high. This suggests that isolated correction of PTC cannot fully determine postoperative shoulder status, and should be matched to that of MTC to achieve optimal PSB.

### Prediction of postoperative CA

In Table 2, the mean TCTA changed from 4.64° to 0.18°, indicating that the corrective surgery largely realigned the thoracic cage. Assuming a rigid connection between the clavicle and the thoracic cage, the post-op CA could be predicted based on the pre-op TCCA. However, in reality, the anatomical connection between the clavicle and the thoracic cage is not particularly rigid, resulting in a significant increase in TCCA postoperatively compared with the pre-op value (from -3.50° to -2.21°). Nevertheless, if this change (ΔCA) can be derived, the predicted post-op CA can still be obtained using the pre-op TCCA (pre-op “true CA”) and ΔCA; specifically, post-op CA = pre-op TCCA + ΔCA. We hypothesize that the factors most closely associated with ΔCA and controllable by the surgeon during the procedure are the correction degree of PTC and the correction degree of MTC. The predictive equation of ΔCA was as follows:

ΔCA = (0.22 × correction degree of PTC) - (0.10 × correction degree of MTC) + 2.86 (R^2^=0.127; correction degree of PTC: B=0.22, *p*<0.001; correction degree of MTC: B=-0.10, *p*=0.009) ⍰

According to the equation ⍰, every 10° of correction in the MTC corresponds to a 1.0° decrease in post-op CA, suggesting that MTC correction has an elevating effect on the shoulder on the concave side. In contrast, every 10° of correction in the PTC corresponds to a 2.2° increase in post-op CA, exerting an opposite effect on shoulder balance; specifically, PTC correction has an elevating effect on the shoulder on the convex side of the MTC.

### Limitations

This study has several limitations. Firstly, this study is image-based, with no data regarding the clinical analysis, such as cosmetic evaluation and patient satisfaction. Secondly, the analysis in this study is limited to immediate postoperative radiographic parameters and thus cannot elucidate the impact of TCCA on postoperative shoulder balance at long-term follow-up. Thirdly, parameters reflecting medial shoulder balance are not investigated in our study. Fourthly, this study is a single-center study, and the number of patients in R group is relatively small. Fifthly, thoracic cage deformity due to spinal deformity may lead to variability in TCTA measurements, but the ICC confirmed that the measurement error was acceptable.

## Conclusions

In Lenke Type 1 and 2 AIS patients, TCCA-based preoperative shoulder balance serves as a direction-specific predictor for immediate PSB. It acts as a significant risk factor for the postoperative ipsilateral shoulder high, while conversely functioning as a significant protective factor against the postoperative contralateral shoulder high. Additionally, the correction rate of MTC is identified as an independent protective factor, and RRCA as an independent risk factor, for postoperative right shoulder high.

## Data Availability

All data produced in the present study are available upon reasonable request to the authors.

